# Factors affecting adoption of COVID-19 prevention measures among community members: A Case study of Entebbe Municipal council in Uganda

**DOI:** 10.1101/2024.05.04.24306856

**Authors:** Eliot Namanya, Nalubega Joy Margret, Nathan Mugenyi, Andrew Twineamatsiko

## Abstract

**Background:** Following continued infections from COVID-19 in Entebbe municipality, there was need to investigate underlying causes off poor adherence. This knowledge would then inform policy and practice enhancements. This study was done to assess the factors affecting the adoption of COVID-19 prevention measures among community members in Entebbe Municipal Council, Uganda given the low levels of adherence to standard operating procedures (SOPs). Specific objectives included to assess the individual factors; the community factors and the social-economic factors affecting the adoption of COVID-19 prevention measures in place among community members in Entebbe Municipal Council, Uganda.

**Methodology:** A descriptive analytical cross-sectional study design was used, employing both quantitative and qualitative methods based on an eligible population of 29,973 and sample of 394 respondents. Primary data was collected using interviews and questionnaires. Qualitative data was analyzed thematically, and quantitative date was analyzed quantitatively using statistical package for social scientists (SPSS) version 23. For quality control, the tool was piloted and found reliable and valid.

**Study Findings:** Results indicate that majority of the respondents were aged 31-40 (32.7%), tertiary level education (76.1%), males (51.5%) and females (48.5%), Christians (84.5%) and Muslims (15.5%), formally employed (78.7%), earned at least a million and above (51.8%). The study explored the factors associated with adoption of Covid-19 SOPs with individual factors, community factors and social economic factors being significant.

**Conclusion:** Individual factors significantly influence adoption of preventive measures. However, the difference between women and men’ adoption and the young people’s negligence to adoption of preventive measures are key.

**Recommendations:** The study recommends subsidizing and/or free distribution of all materials and equipment that prevent communicable diseases; use influential members of the public as health ambassadors to influence positively the public; free protective gears to groups of people who frequently interact with communities; improved transport system.

## Introduction

Several economic and social disruptions have existed both in Uganda and globally owing to the invasion of the Corona virus which caused COVID-19 (Auriemma and Iannaccone, 2020) The World Health Organization (WHO) noted that we have a long way to such disruptions since the virus will be with us for a long time (WHO, 2020). This makes adherence to the standard operating procedures vital as there are gaps in adherence whose associated factors are yet to be understood in the Ugandan context (Lippke *et al*., 2022)

China reported the first case of Covid-19 in Wuhan in December 2019 which then spread to all parts of the world including African countries (Luo *et al*., 2020). The World Health Organization (WHO) acknowledged COVID-19 to be a public health emergence of international concern (PHEIC) on January 30^th^, 2020 and after this it spread widely to become a global health concern (Nadeem *et al*., 2020). COVID-19 is an illness caused by a novel corona virus that is Severe Acute Respiratory Syndrome (SARS) corona virus (Hui-jun *et al*., 2020). There are several signs of COVID-19 including fever, cough, fatigue and dyspnea symptoms while in the severe forms, it causes severe acute respiratory distress (Hu *et al*., 2020).

The mode of transmission of COVID-19 includes the respiratory secretions which is the most likely source of transmission and commonest cause of viral transmission. Other sources include contact with contaminated surfaces since a virus remains viable for eight hours on them (Kampf *et al*., 2020). Co-morbidities such diabetes, hypertension, malignancy, and cardiovascular diseases increase the risk of succumbing to COVID-19 (Bloomgarden, 2020). For instance, the pooled estimates of critical cases in diabetes and hypertension are high (Hu *et al*., 2020). Persons living with HIV that also have other underlying co-morbidities and are at a great disadvantage from the negative consequences of COVID-19. Thus, it becomes very important for such persons to comply with the preventive measures (Waterfield *et al*., 2021). Using previous experience in handling pandemics as documented by (Parpia *et al*., 2016), Africans leaders took action to contain covid-19 and its causing agent the Corona-Virus. Uganda is also not exception, having registered her first COVID-19 positive case on the 31^st^ of March 2020 (Kiwanuka *et al*., 2020).

Globally, public health measures like face masks, hand hygiene and maintaining social distancing have been implemented to delay and reduce local transmission of COVID-19 (Abdullahi *et al*., 2020). Several guidelines have been put in place as the disease has been progressing and these guidelines are regularly updated as knowledge on COVID-19 expands (WHO, 2020). In Thailand, the prevalence of face mask utilization was estimated at 46%. In Paris, the capital of France, out of 356 observations, there was only 4% of those observed wearing facemasks. In the US, in the cities of Boston and Atlanta only 3% and 2% of 371 and 554 observations were wearing facemasks respectively. In Lima, the capital of Peru, 27% of 237 observations worn facemasks (Elachola, Ebrahim and Gozzer, 2020). South Korea, Vietnam, and Taiwan are considered as exemplary around the world in their public health response to COVID-19 (Shokoohi, Osooli and Stranges, 2020). This shows that the rate of wearing face masks globally including developed countries is low which increases risk of spreading COVID-19. In China, adoption of facemask wearing in public places was 95.7%; sanitizing hands regularly after returning from public spaces was 70.9% adoption (Pan *et al*., 2020).

To combat the spread of COVID-19, several SOPs were developed and the public was encouraged to adhere to them (Onigbinde *et al*., 2021). Among these included; avoid public transportation; minimize contact with passengers, minimize going out into the general population; avoid crowds, shopping malls, sporting or mass events, and other situations likely to attract large numbers of people; wash your hands often with soap and water for at least 20 seconds or use at least a 60% alcohol based hand sanitizer; avoid touching your face eyes, nose and mouth (World Food Programme, 2021).

In Africa, physical distancing measures have made it possible to contain the spread of COVID-19 like in Western countries without too much questioning about their acceptability of the SOPs. Whereas SOPs are generally good, and despite their overall acceptance, they have been poorly documented, particularly in Africa. Many political, ethical and socioeconomic questions arise (Ahmed *et al*., 2020). In Mali, an international religious gathering, with thousands of participants, was able to take place in the region of Sikasso. In Burkina Faso, prayers in places of worship were not immediately banned and markets remained open for a long time (Ahmed *et al*., 2020). In Guinea, the curfew was lifted for towns in the interior of the country (Dixon *et al*., 2020). Here, we see that social-demographic factors, notably, religion affected compliance with the SOPs.

In Uganda, the initial national wide COVD-19 prevention measure undertaken by MOH was a countrywide lockdown that was initiated on the 30^th^ March 2020 (Kiwanuka *et al*., 2020). Then later other preventive measures including facemasks, social/physical distancing, regular-proper hand washing with a disinfectant among others were put in place (WHO, 2020). This reduced on the COVID-19 cases in Uganda. Besides the SOPs, Uganda put the following measures in place; Closing of educational institutions, Banned Ugandans from moving to or through category one (I) countries that had had a large number of corona cases by that time; Burials with 10 people maximum; no weekly or monthly markets, Suspended all the discos, dances, bars, sports, music shows, cinemas and concerts; Advised the public on good nutrition to strengthen the body defense system (Africa CDC, 2019). The number of COVID-19 cases remained low between March and June 2020; however, when these measures were relaxed towards the end of June 2020, there was an increase by 88% in the number of new cases reported each week where majority were from Kampala (Africa CDC, 2019). Limited compliance with SOPs was associated with decreasing compliance with SOPs although no empirical evidence was provided hence a knowledge gap.

While the practice of hand washing was possible and adhered to at the beginning of March, it has radically dropped by up to 92.6% in Kabalagala and Kimombasa (Odoi, 2020). The police to Division leaders, food distributors, VHTs and local village leaders were not working in harmony and were giving different, sometimes confusing, messages and instructions. This greatly contributed to confusion and partly resulted in community non-compliance. In Uganda, communities have had multiple - and sometimes contradictory - sources of information which affect their adherence to the preventive measures (Odoi, 2020).

This necessitates a study to assess the factors that affect the adoption of COVID-19 prevention measures among the communities in Uganda since it is the means to reducing transmission. To have a close focus, the current study covers the adherence and related factors in Entebbe municipality. Most studies have however focused on how the money for covid-19 was spent, how the task force performed, how covid-19 has interrupted business and the economy and so on (Muhwezi *et al*., 2020). There is a gap in knowledge on how the community has reacted and perceived the SOPs in Uganda.

In Entebbe municipality, Mandela reports the mayor being concerned about children whose need for socialization made them play and loose masks as well as forgetting about social distancing. The adults often neglect the guidelines and instead look for survival and in the process losing touch with most of the SOPs (Mandela, 2020). As a result, an increase in the COVID-19 cases led to closure of the hospital for other services apart from emergencies and COVID-19 cases in late 2020 and early 2021 (Adude, 2021). All these incidences make Entebbe a special case for investigation of adherence to the SOPs and the future containment of the covid-19 virus spread. What is known is that Entebbe remains one of the hotspots for COVID-19, however, little is known through proper documentation about the adherence to the SOPs and the associated factors. With daily admissions of 10 COVID-19 patients for months (Adude, 2021), clearly, there are gaps in adherence that need to be accounted for (Kadowa, 2020), reports that the control measures put in place have slowed the spread of the pandemic, have delayed widespread community infection and contributed to the few reported deaths so far. A Lancet Commission report presented at the 75th United Nations General Assembly ranked Uganda among the ten countries that had achieved suppression of the pandemic in August 2020. The author however does not show details of embedded factors.

## Conceptual Framework

This study is guided by the Health Belief Model (HBM). The HBM model can be used to predict and explain influencing behaviors (Rosenstock, Strecher and Becker, 1988). This informed the choice of this theory for our study. The HBM model was used while attempting to show how different variables may be related, and in so doing, we posited a conceptual explanatory model for our study. According to the HBM model, traditional health belief variables of perceived susceptibility, severity, benefits and barriers, incentives to behave (health motivation) and locus of control influence and predict an individual’s response to a disease or disease-causing agents (Rosenstock, Strecher and Becker, 1988). Perceived susceptibility has been well established as a major force driving adherence to facemask-wearing, with a higher perception of susceptibility being linked to higher compliance with mask-wearing. Persons who personally fell very vulnerable to contracting COVID-19 are 2.5 times more likely to wear facemasks. In terms of psychological factors, perceived susceptibility to becoming infected by the virus is usually considered to increase the likelihood of compliance with social distance behaviors (Yilmazkuday, 2021).

Utilization of prevention measures for every disease condition including COVID-19 are motivated by the person’s perceived risk susceptibility as well as the benefits of taking such prevention measures and such key elements of the Health Belief Model can be used to design or adapt health promotion or disease prevention programs.

## Materials and Methods

### Research Design

A cross-sectional study design was used, employing both quantitative and qualitative methods. This design was chosen as it allows for description of variables and assesses their association on the community under study (Aggarwal and Ranganathan, 2019).

(Cresswell and Plano Clark, 2011) define mixed-methods research as those studies that include at least one quantitative strand and one qualitative strand. A strand is a component of a study that encompasses the basic process of conducting quantitative or qualitative research: posing a research question, collecting, and analyzing data, and interpreting the results.

(Cresswell and Plano Clark, 2011) adds that, qualitative data provide understanding through greater depth, whereas quantitative data provide broader, more general understanding. Each approach has its advantages and limitations. Qualitative data may provide a deep examination of a phenomenon of interest but only with respect to a handful of participants. On the other hand, quantitative data can provide information across a much broader sampling of participants, but the depth of that information is certainly limited. Using both methods checks limitations of each of them to get an enhanced report.

### Area of Study

Entebbe municipality is located on the shores of Lake Victoria; it has a total area of 56 Square kilometers (sq.km) with 20 sq.km covered by water Lake Victoria waters. The coordinates of Entebbe municipality are 0°03’00.0"N, 32°27’36.0"E (Latitude: 0.0500; longitude: 32.4600), it is located 34 km southwest of Kampala capital city. Entebbe Municipality is one of the areas with low poverty levels in Uganda. However, there are certain arrears that are specifically hurt by poverty namely, Lwamunyu landing sites, Musoli, in Kigungu ward, Lugonjo-Nakiwogo and Kitoro Central in Kwafu ward and Katabi-Busambaga in Katabi ward.

The study was carried out in Entebbe municipality in Wakiso district. The data was collected from all the cells and villages comprising Entebbe municipality and randomly between April and July 2022. This was motivated by the poor adherence to the covid-19 preventive measures up to more than 10000 in a just a few months on the onset of Covid-19. The study area was selected because it has the high number of Covid-19 cases in all the regions in Kampala metropolitan area and also with a good mix of people of various social economic backgrounds in terms of tribe, religion, beliefs, income, occupations among others making it a suitable area to carry out the investigation. It has 62,969 people (Uganda Bureau of Statistics, 2016). Entebbe Municipality has civil servants, contractors, casual laborers, pensioners, artisan, brick makers, vehicle repairers, fisher folk, farmers, traders, hoteliers and aviation related occupations.

### Source Population

The population in this study constitutes all persons working, residing, and commuting through Entebbe municipal council on a regular basis. Whilst the target population constitutes residents of Entebbe Municipal council residing in this area during the period when this study was conducted. Entebbe municipality has a population of 62,969 of which 29973 are above 18 years (2017).

### Study Population

According to the latest national census (Uganda Bureau of Statistics, 2016), people less than 18 Years old constitute 52.6%. If this is applied to Entebbe municipality, the study population comprised (47.4/100) * 62,969. This resulted into a population of 29973 that were eligible for the study.

### Sampling procedures

#### Simple random sampling

For general respondents other than the leaders, the study used Simple random sampling (SRS). This involved giving the questionnaire to any accessible and consenting adult within Entebbe municipality. SRS is advantageous for surveys at the community level; it allows equal chances and guarantees the researcher to get to get a representative sample of the sampling frame (Polit and Beck, 2008). Based on listings of community households from Entebbe Municipality, one person who fills the inclusion criteria from each selected household who participated in the study.

#### Purposive sampling

For leaders, purposive was used. Here, only leaders were selected for interviews to ensure that they give a detailed account of covid-19 SOPs adherence status and underlying factors (Palinkas *et al*., 2015).

#### Sample size calculation

Slovin’s formula was used to calculate the sample size necessary to achieve a certain confidence interval when sampling a population.

Slovin’s formula is written as: n = N/(1+Ne^2^),

Where: n= the number of samples N=the total population n=sample

N=population (29973)

e= error (0.05)

A total of 394 respondents was considered.

### Data collection techniques/methods

#### Questionnaire (quantitative method)

The questionnaires were given physically to the respondents and in some cases, online email survey was used, and results merged later for analysis. The study used a questionnaire to collect quantitative data for each of the study variables.

#### Interview guide (qualitative method)

An interview guide was used to conduct in-depth interviews from the leaders in Entebbe municipality. With an interview, detailed information was easily captured.

### Data collection tools

#### Interview Guide

The study designed an interview guide in line with the study objectives. The interview guide was used to collect data from the leaders, notably, political leaders, religious leaders, opinion lead leaders who were part of the study. Data collected were analyzed thematically.

### Quantitative Tool

#### Self-Administered Questionnaires

The study used a structured self-administered questionnaires to collect data. This method limited the physical interaction of respondents and in view of Covid-19; this was a precautionary measure yet enable data collection as planned. The questionnaire was given to all respondents groups apart from those identified for interviews. A tape recorder was used to provide unbiased information that was analyzed later.

#### Quality control techniques

Quality control in this study aimed at reducing errors, redundancies, inconsistencies and ensured correct and complete information is obtained. To achieve this, the study piloted tools and conducted validity testing and reliability testing.

#### Validity for qualitative data

Triangulation, which is using two or more methods to study the same phenomenon, was used. The study triangulated the tools only. The study was introduced in a manner that allowed participants adequate time and ability to freely consider whether or not they wish to take part. This further ensured that data was collected from valid individuals as suggested by (Sarantakos, 2007).

#### Validity for quantitative data

This was used to check all items in the tools to ensure that all the questions are valid. To ensure validity, all the data collection tools were evaluated by two experts independently. These rated every question as relevant, somewhat relevant and irrelevant. Two experts were requested to rate tool and the threshold was 0.7, then the tool would be considered acceptable, or else it was re-designed to suit the relevancy scale (Sekaran and Bougie, 2016).

#### Reliability for qualitative data

For the qualitative results, reliability was ascertained by comparing the results obtained with other evidence and making observations more than once and ensuring that informants were explained clearly the nature of the research (Mugenda, O. M., 2003).

#### Reliability for quantitative data

Before the questionnaire was used in the real study a pilot study on respondents from another location was done to determine their reliability (Saunders, Lewis and Thornhill, 2003). This pre-test comprised a sample of all respondent categories with similar characteristics but from another area. To determine the reliability of the questionnaire, SPSS software version 23 was used. This followed the procedure of analyse, reliability, scale and the cronbach’s alpha coefficients were generated, with a minimum score of 0.7, to pass the reliability test.

From the table above, result confirm that the variables met the validity and reliability threshold of 0.7 as the minimum acceptable score for both validity and reliability (Saunders, Lewis and Thornhill, 2003).

### Data analysis

#### Quantitative data analysis

Quantitative data analysis was done using IBM SPSS software 23. Univariate descriptive statistics including frequencies, means and standard deviations were done to reveal sample characteristics. Bivariate analysis was done to infer on the correlates between variables, inferential statistics using Chi-square test, analysis of variance (ANOVA) and logistics regression analyses was done.

#### Qualitative data analysis

Thematic analysis was used for qualitative data.

#### Ethical considerations

Ethical clearance was sought from the Committee on Research Ethics. Clearance to collect data from the setting was also sought from the administration of the study settings to conduct the study. An informed consent was used before data collection.

## Results

### Response rate

The response rate was 100%

### Respondents demographics

The study explored the demographics of the respondents and the results obtained are presented in table 3 below.

**Table 1:**
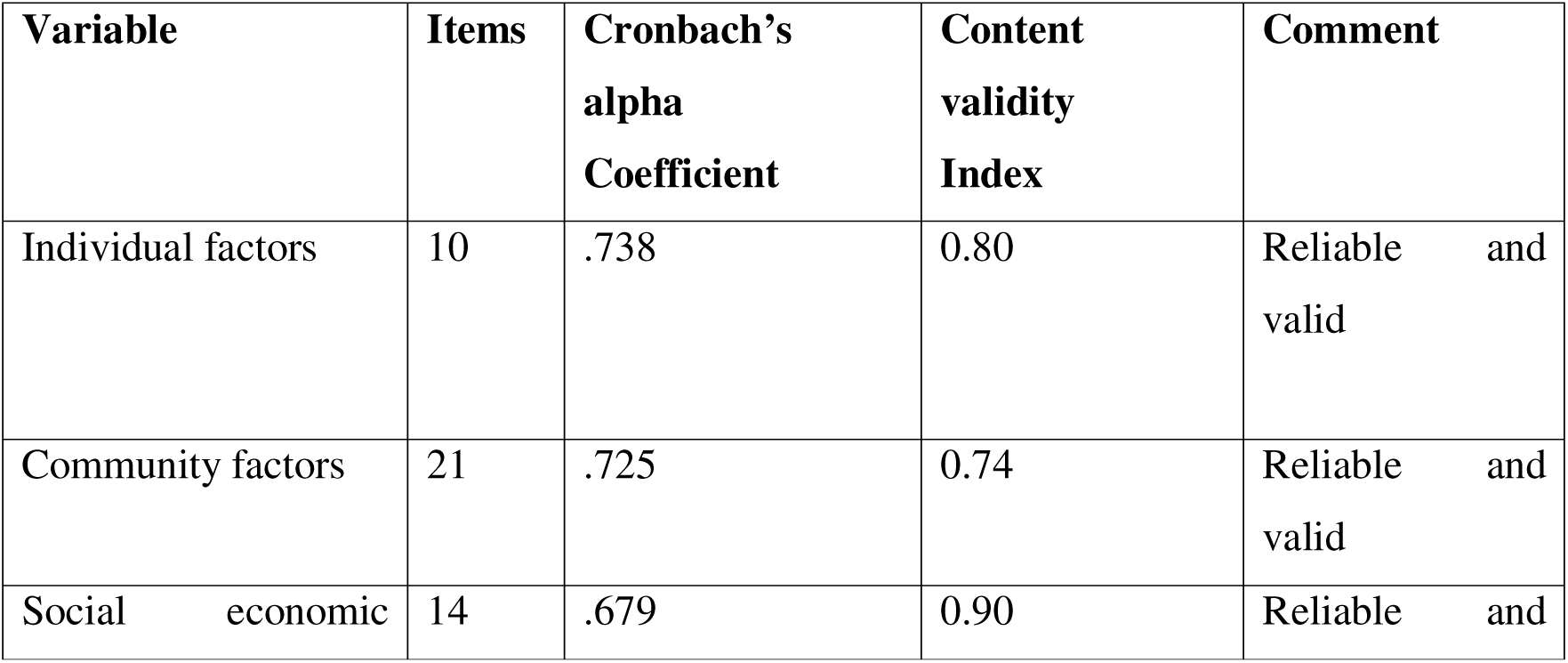
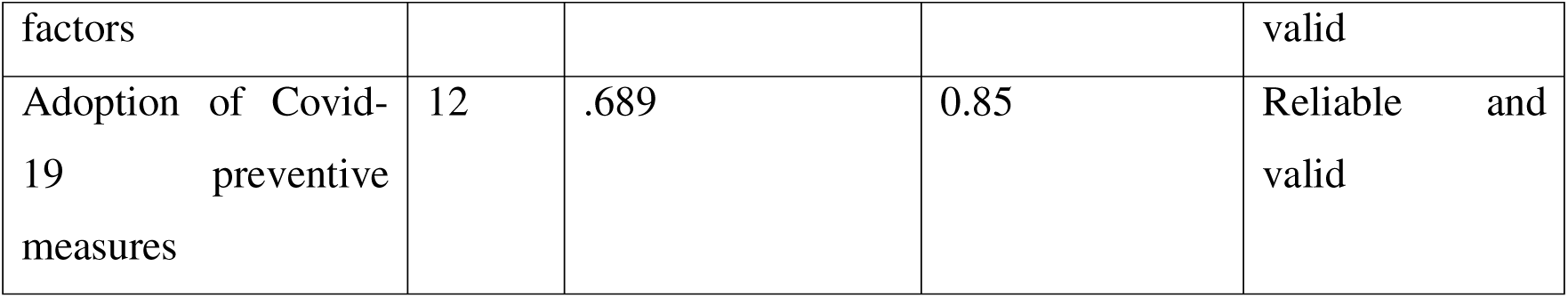
Validity and reliability statistics.

**Table 2:** Age, marital status, level of education, gender.

| **Age In Years** | **Frequency** | **Percent** |
| --- | --- | --- |
| 18-20 | 13 | 3.3 |
| 21-30 | 75 | 19.0 |
| 31-40 | 129 | 32.7 |
| 41-50 | 107 | 27.2 |
| Above 50 | 70 | 17.8 |
| **What is your marital status?** | Frequency | Percent |
| Cohabiting | 46 | 11.7 |
| Married | 267 | 67.8 |
| Single | 74 | 18.8 |
| Widow | 4 | 1.0 |
| Widower | 3 | 0.8 |
| **What is your level of education?** | Frequency | Percent |
| BNS | 1 | 0.3 |
| Masters | 3 | 0.8 |
| Post graduate diploma | 1 | 0.3 |
| Primary | 12 | 3.0 |
| Records personnel | 1 | 0.3 |
| Secondary | 71 | 18.0 |
| Tertiary | 300 | 76.1 |
| Undergraduate | 1 | 0.3 |
| University | 3 | 0.8 |
| YMCA comprehensive | 1 | 0.3 |
| **What is your gender?** | Frequency | Percent |
| Female | 191 | 48.5 |
| Male | 203 | 51.5 |
| Total | 394 | 100.0 |
| **What is your religion?** | Frequency | Percent |
| Christian | 333 | 84.5 |
| Muslim | 61 | 15.5 |
| **What is your tribe?** | Frequency | Percent |
| Acholi | 14 | 3.6 |
| European | 1 | 0.3 |
| Gisu | 18 | 4.6 |
| Iteso | 6 | 1.5 |
| Kenyan | 2 | 0.5 |
| Kiga | 32 | 8.1 |
| Kojo | 3 | 0.8 |
| Langi | 4 | 1.0 |
| Lugbara | 5 | 1.3 |
| Luo | 1 | 0.3 |
| Mixed | 1 | 0.3 |
| Mufumbira | 4 | 1.0 |
| Muganda | 191 | 48.5 |
| Munyankore | 41 | 10.4 |
| Munyole | 1 | 0.3 |
| Nubian | 1 | 0.3 |
| Nyole | 1 | 0.3 |
| Nyoro | 4 | 1.0 |
| Omuhororo | 1 | 0.3 |
| Rwandese | 1 | 0.3 |
| Samia | 1 | 0.3 |
| Soga | 30 | 7.6 |
| Somali | 3 | 0.8 |
| Sundanese | 3 | 0.8 |
| Toro | 25 | 6.3 |
| What is the nature of your employment | Frequency | Percent |
| Formal | 310 | 78.7 |
| Informal | 56 | 14.2 |
| None | 25 | 6.3 |
| Other | 3 | 0.8 |
| What is your monthly salary? | Frequency | Percent |
| <50000/= | 22 | 5.6 |
| 50001 - 100000/= | 14 | 3.6 |
| 100001 - 500000/= | 33 | 8.4 |
| 500001 - 1000000/= | 121 | 30.7 |
| Above 1000000/= | 204 | 51.8 |
| Total | 394 | 100.0 |

**Table 3:**
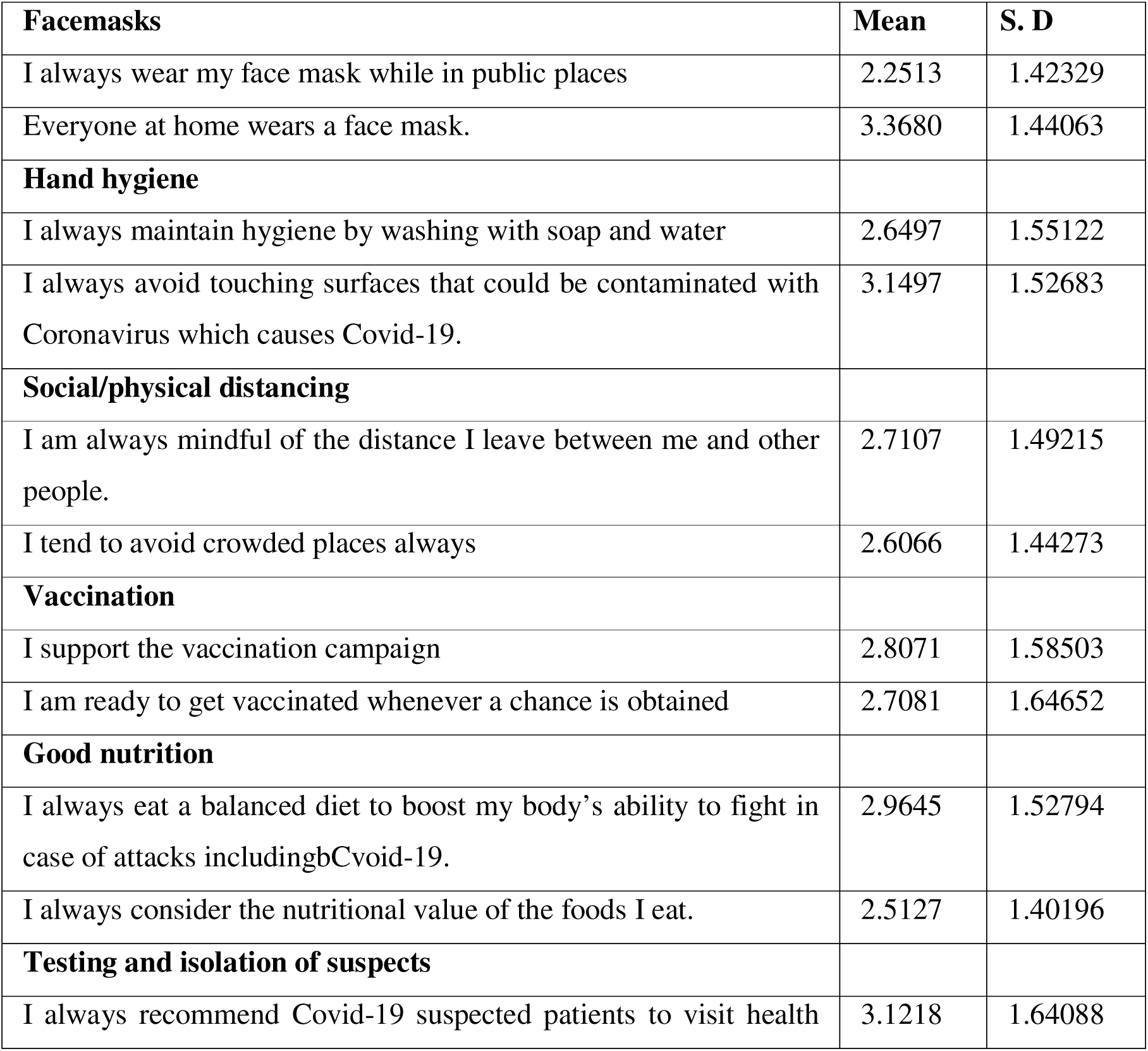
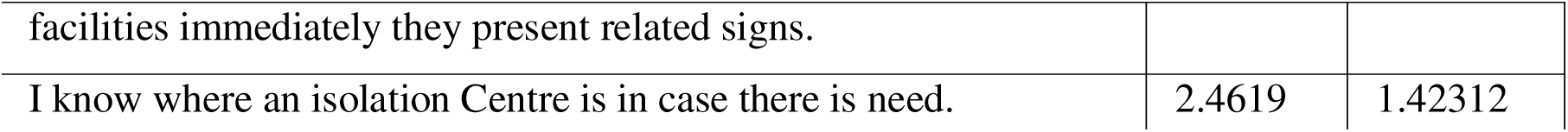
Adoption of COVID-19 Prevention Measures.

Results indicate that majority of the respondents were aged 31-40 (32.7%) followed by those aged 41-50 (27.2%) indicating that majority of the participants were mature people with genuine concern for Covid-19 and related issues.

Regarding marital status, results indicate that majority were married 267(67.8%). The singles and cohabiting groups were also well represented by 18.8% and 11.7% respectively with 1% and 0.8% representing the widow and widower groups respectively. This means that Covid-19 was a concern of all people regardless of their marital status.

Results on education reveal that the tertiary level took the lead with 300 (76.1%) followed by secondary 71(18%) among other groups. These results confirm that participants largely were literate enough to give reliable information and understood the tool’s language. Again, it shows that Covid-19 prevention was a concern of both the highly educated and those with less education.

Results on gender reveal more male participation at 203(51.5%) compared to female participation at 191(48.5%). This confirms that men and women are equally concerned about Covid-19 issues hence participation in almost equitable tables.

### Employment and salary

From table 4.1, results confirm that majority of the respondents were formally employed 310 (78.7%). This means that an average adult in Entebbe is formally employed and thus has an income, regardless of how much is earned.

Being an urban center next to the airport, the results reflect that most respondents were earning at least a million and above, represented by 204(51.8%), followed by those earning between 500,001 and 1,000,000. This shows that an average person in Entebbe earns monthly 500,000 and above. However, given the cost of leaving where an average expenditure of a family of 6 spends 1.5 million shillings on only food and accommodation (as revealed from interviews), this income is not so impressive. However, having some income makes its more possible to buy SOPs related equipment like sanitizer, masks, temperature guns and so on.

### Adoption of COVID-19 Prevention Measures among Community Members in Entebbe Municipal Council, Uganda

The results on the adoption of COVID-19 Prevention Measures among Community Members in Entebbe Municipal Council, Uganda are presented in table 4 below.

**Table 4:**
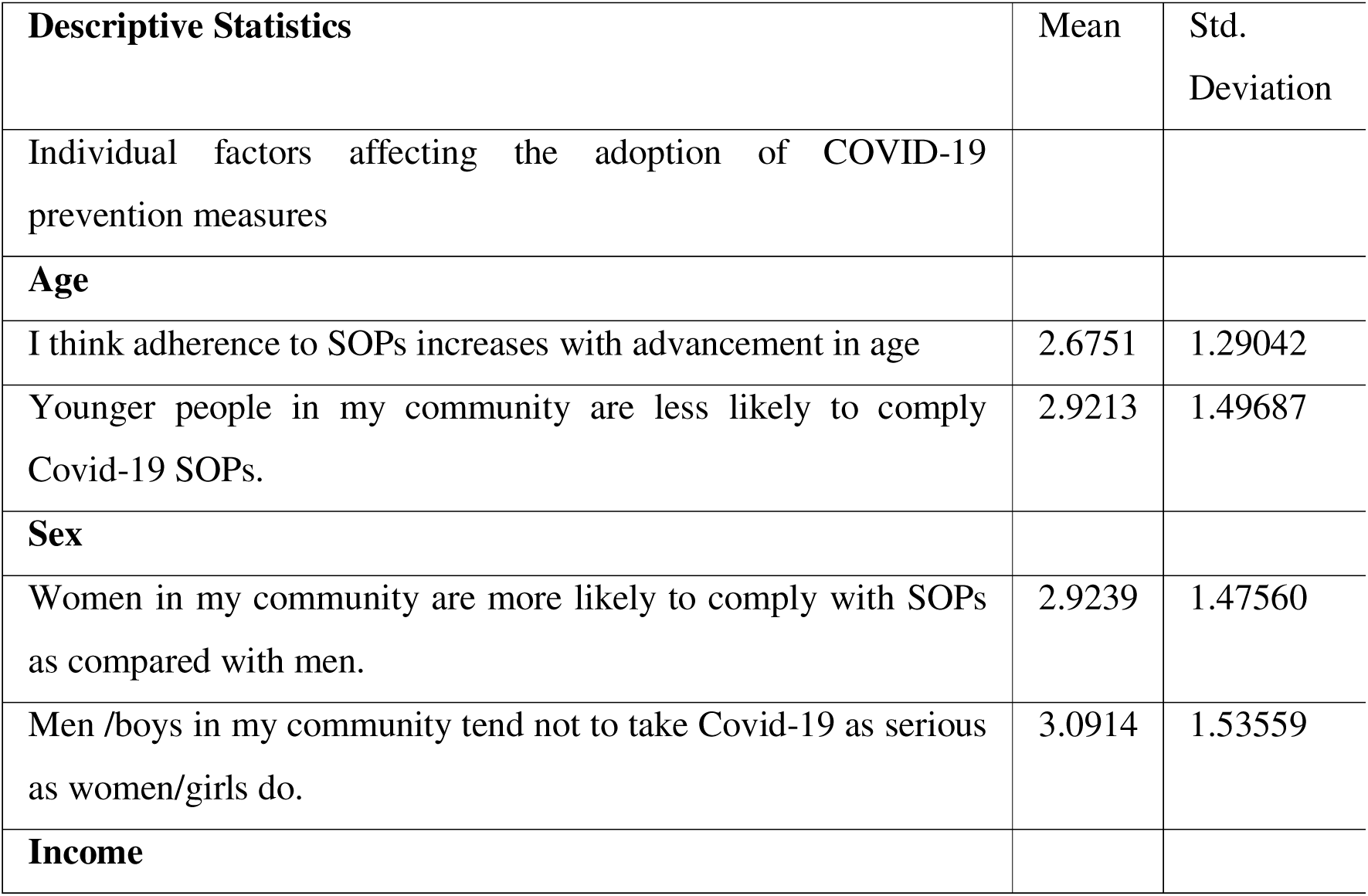
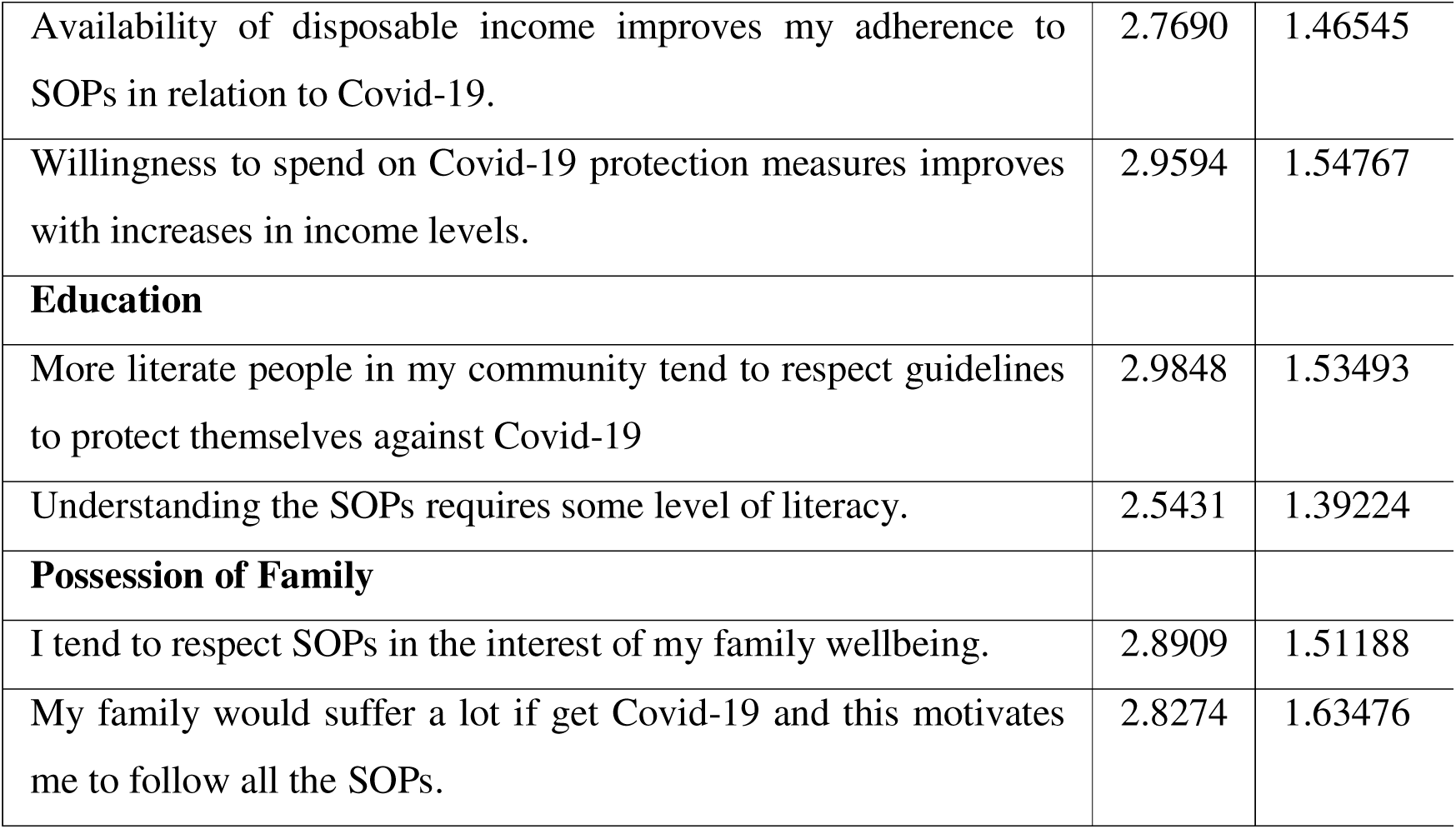
Individual Factors Affecting the Adoption of COVID-19 Prevention Measures.

**Table 5:** Correlation between Individual Factors Affecting and Adoption of COVID-19 Prevention Measures among Community Members in Entebbe Municipal Council, Uganda.

| **Correlations** |  |  |  |  |  |  |  |
| --- | --- | --- | --- | --- | --- | --- | --- |
| **Variable** | **1** | **2** | **3** | **4** | **5** | **6** | **7** |
| Age (1) | 1 |  |  |  |  |  |  |
| Sex (2) | .035 | 1 |  |  |  |  |  |
| Income (3) | .150<sup>**</sup> | .041 | 1 |  |  |  |  |
| Education (4) | .062 | .160<sup>**</sup> | .112<sup>*</sup> | 1 |  |  |  |
| Family (5) | .032 | .196<sup>**</sup> | .249<sup>**</sup> | .218<sup>**</sup> | 1 |  |  |
| Individual factors (6) | .429<sup>**</sup> | .520<sup>**</sup> | .557<sup>**</sup> | .568<sup>**</sup> | .661<sup>**</sup> | 1 |  |
| Adoption (7) | -.017 | .160<sup>**</sup> | .229<sup>**</sup> | .285<sup>**</sup> | .428<sup>**</sup> | .412<sup>**</sup> | 1 |
| **. Correlation is significant at the 0.01 level (2-tailed). |  |  |  |  |  |  |  |
| *. Correlation is significant at the 0.05 level (2-tailed). |  |  |  |  |  |  |  |

**Table 6:** Regression between Individual Factors Affecting and Adoption of COVID-19 Prevention Measures among Community Members in Entebbe Municipal Council, Uganda.

| Model Summary |  |  |  |  |  |  |  |
| --- | --- | --- | --- | --- | --- | --- | --- |
| Model |  | R | R Square |  | Adjusted R Square |  | Std. Error of the Estimate |
| 1 |  | .412<sup>a</sup> | .170 |  | .167 |  | .65584 |
| a. Predictors: (Constant), Individual factors |  |  |  |  |  |  |  |
| ANOVA<sup>a</sup> |  |  |  |  |  |  |  |
| Model |  | Sum of Squares | Df | Mean Square | F | Sig. |  |
| 1 | Regression | 34.424 | 1 | 34.424 | 80.032 | .000<sup>b</sup> |  |
|  | Residual | 168.608 | 392 | .430 |  |  |  |
|  | Total | 203.032 | 393 |  |  |  |  |
| a. Dependent Variable: adoption |  |  |  |  |  |  |  |
| b. Predictors: (Constant), Individual factors |  |  |  |  |  |  |  |
| Coefficients<sup>a</sup> |  |  |  |  |  |  |  |
| Model |  | Unstandardized Coefficients |  | Standardized Coefficients | t | Sig. |  |
|  |  | B | Std. Error | Beta |  |  |  |
| 1 | (Constant) | 1.280 | .170 |  | 7.508 | .000 |  |
|  | Individual factors | .523 | .059 | .412 | 8.946 | .000 |  |
| a. Dependent Variable: adoption |  |  |  |  |  |  |  |

### Overall score on adoption of COVID-19 Prevention Measures

Results in table 4 show that the overall mean on adoption was 2.863 and standard deviation of 1.496 This means that largely respondents were not sure of the level of adoption of the Covid-19 preventive measures. This was also generated with a high degree of deviation thus a sharp difference in the opinions of the different participants.

### Facemasks

Face masks as a measure of prevention scored 2.810 as mean and standard deviation of 1.432. This means there were doubts on the level of usage of face masks while significant differences in opinions on the same are seen as revealed by the high standard deviation.

### Hand hygiene

Hand hygiene scored a mean of 2.900 and standard deviation of 1.539. This means that hand hygiene usage was not ascertained and there were sharp contrasts in the opinions given on this.

### Social/physical distancing

Again, social distancing with a mean score of 2.855 and standard deviation of 1.485, reveal that respondents still were not sure of the level of adherence on this with contrasting opinions as revealed by the high standard deviation.

### Vaccination

Still, vaccination scored a mean of 2.877 and standard deviation of 1.512, meaning that majority were not sure of the level of vaccination attained with a sharp contrast in the opinions given the high standard deviation.

### Good nutrition

Good nutrition scored a mean of 2.866 and standard deviation of 1.499. This means that where good nutrition is recommended, there were differences in the level of adherence and contrasting opinions on the same among respondents.

### Testing and isolation of suspects

The mean score on testing and isolation was 2.872 and standard deviation of 1.506. This means that still, there were divergent opinions on the same hence not being certain overall.

### Highest and lowest scores

In absolute terms, the most impressive scores were recorded on respondents wearing a face mask while in public places (mean =2.2513, SD=1.423) and knowing where an isolation Centre was in case there was need (mean =2.4619, SD=1.42312).

Least impressive scores were obtained on always avoiding touching surfaces that could be contaminated with Coronavirus which causes Covid-19 where 190 agreed (48%) and 194 disagreed (49%). and that everyone at home was wearing a face mask, where 132 agreed (%) and 236 disagreed (60%).

### Qualitative Results

For qualitative results, the following pseudo names have been adopted; political leaders= PL; religious leaders= RL; opinion lead leaders= OP. This applies to all the qualitative results in this report. The interviews carried out with political leaders, religious leaders and opinion leaders had some revelations as to the levels of adoption of COVID-19 prevention measures as described below.

### Theme 1: personal initiatives

#### a) Facemasks

Face masks were generally accepted and used by the community in Entebbe.

> “*Our people are now getting used to masks, actually just follow our example as leaders with no questions” (PL2, Male, 41-45)*

In support of the same idea, one of the opinion leaders proudly noted,

> *“In my zone, both elders and young people follow my ideas. When Covid-19 came, I appealed to them to put on masks and since then, they wait for my other instruction “(OP2, Female, 51-55)*.

In the above examples, selected out of the various responses from the interviews, there is confirmation that masks were increasingly being adopted as a preventive measure for Covid-19 in Entebbe.

#### b) Hand hygiene

Hygiene, especially in relation to hand washing using soap and water and use of sanitizers were increasingly adopted especially by women. This was revealed in interviews with a few examples cited here as representatives of the views obtained,

> *“Most women here who are the custodians of family hygiene had fully embraced hand washing. We are actually fighting not only Covid-19 but also other diseases like diarrhea, dysentery among others” (RL1, Female, Christian 56-60)*

It was also revealed that for the Muslim community hand washing was already the norm as indicated here,

> *“as a religious leader, I do not only leady Muslims since God is for everyone, whoever, hand washing, and general hygiene are part of the Muslim community and I see Christians also following up. With the help of Allah, we shall overcome this” (RL2, Male, Moslem 51-55)*.

For political leaders, Covid had some advantages and politicians used this as a chance to show off their care for the community, as highlighted here,

> *“When Covid came, we were all afraid since we are human. However, I have added my contribution by donating hand washing equipment’s to many zones. I have secured political capital and at the same timed been helpful in ensuring that people wash, and I believe, they are washing hands seriously.” (PL3, Female, 36-40)*.

### Theme 2: community initiatives

#### a) Social/physical distancing

All respondents including religious leaders, politicians and opinion leaders agreed that social distance was poor. This was connected to poor housing, the need to economize on public transport costs, events that abruptly attracted gathering like people getting accidents, arresting thieves, burials and so on. On this note, this had failed miserably. However, the use of face masks was noted to reduce the risks that would follow disrespect for social distancing. One of the leaders noted how this had failed,

> *“In Entebbe people try to respect social distancing. However, if any event occurs like burial, or even coming of a leader, people forget all about social distancing. Fortunately, many have masks” (PL1, Male, 41-45)*.

#### b) Vaccination

Vaccination was in high gear, however, being surrounded by myths and misinformation. Most leaders lamented that government had not done enough to sensitize people how safe the vaccine was and even those who took it up had double thoughts, without being sure.

> *“Personally, I am not a scientist, but I believe the vaccines are safe. However, it takes time for me to convince people that the vaccine is safe, and government only run adverts randomly without proving if the target audience is listening.” (PL3, Female, 36-40)*

Similar or related views were obtained from other leaders. The general impression is that people took the vaccine although not much was done to give then adequate information on platforms they liked most and, in a form, they could understand and from people they would trust.

#### c) Good nutrition

It was noted that people easily understood and followed good nutrition except for those who were financially constrained. Leaders and opinion leaders all agreed that eating well was even cheaper than eating badly. Society was able to appreciate this as it was seen to be totally safe. As such, most families had added fruits like oranges to their daily menu to boost immunity. This was the impression obtained from all the interviewed respondents.

#### d) Testing and isolation of suspects

Testing and isolation of suspects was more of a medical process and the leaders interviewed did not give much on this. However, they encouraged whoever had common symptoms of Covid-19 to go the nearest health facility as quickly as possible.

### 4.4 Individual Factors Affecting the Adoption of COVID-19 Prevention Measures among Community Members in Entebbe Municipal Council, Uganda

The study explored individual factors affecting adoption of Covid-19 Prevention Measures among Community Members in Entebbe Municipal Council, Uganda. The results obtained on this are presented in table 7 below.

**Table 7:** Community Factors Affecting Adoption of COVID-19 Prevention Measures.

| **Descriptive Statistics** | **Mean** | **Std. Dev** |
| --- | --- | --- |
| **Sensitization modes** |  |  |
| Use of electronic media like social media, TV among others has been helpful in improving my adherence to SOPs. | 2.7183 | 1.62497 |
| The use of non-electronic media like radios, newspapers, community public address etc. has proved more effective in improving my adherence to SOPs. | 3.0660 | 1.55172 |
| There is sensitization strategy in my community. | 2.4848 | 1.36515 |
| The methods used to relay information to my community in relation to SOPs have done a great job to improve SOPs adherence. | 3.1878 | 1.53177 |
| **Settlement pattern** |  |  |
| Being in a crowded place interferes with adherence to SOPs. | 2.2284 | 1.33598 |
| The rural community is better able to follow SOPs as compared to the urban population | 3.2893 | 1.50234 |
| Our house has adequate rooms to ensure adherence to SOPs like social distancing. | 2.9569 | 1.57288 |
| People who live in their own houses (not renting) are more likely to follow the SOPs compared to those in congested rented houses. | 3.2081 | 1.55553 |
| **Peer influence** |  |  |
| The youth in the community tend to follow their peers who do not adhere to the good practices | 2.6396 | 1.42213 |
| Members who share common trade e.g. market workers, taxi drivers tend to be influenced by each other to either or not to adhere. | 3.2919 | 1.58350 |
| People who abuse drugs tend to influence people near them not to adhere to SOPs. | 2.7030 | 1.43410 |
| **Perceptions** |  |  |
| It is discomfort able to use the covid-19 prevention measures. | 2.8807 | 1.46841 |
| The community thinks that Covid-19 is not purely a health issue. | 3.2386 | 1.53796 |
| People who have not lost close friends or relatives do not take SOPs seriously. | 2.8756 | 1.63370 |
| **Safety of face masks** |  |  |
| I think wearing face masks is important. | 2.5812 | 1.56610 |
| Most people like to wear face masks properly | 3.2614 | 1.52842 |
| I think most of the face masks in place are effective. | 3.0051 | 1.54491 |
| **Risk perceptions** |  |  |
| I have chronic illness that hinder me from adopting to covid-19 prevention measures | 3.4162 | 1.50465 |
| I know Covid-19 is potentially fatal. | 2.7640 | 1.50068 |
| I have lost people known to me due to Covid-19. | 3.0381 | 1.58751 |
| I know people to who are currently suffering from Covid-19. | 2.7640 | 1.50576 |

### Overall score on Individual Factors

Results reveal a mean score of 2.919 and standard deviation of 1.458. This shows that overall, there was no certainty on how individual factors influence adoption of Covid-19 prevention measures.

### Age

Age scored a mean of 2.798 and standard deviation of 1.394. This means that participants were not sure if age affected adherence to Covid-19 preventive measures.

### Sex

Sex scored a mean of 3.008 and standard deviation of 1.506. This means that means that there was uncertainty on whether sex affected adherence to Covid-19 preventive measures.

### Income

Income scored a mean of 2.903 and standard deviation of 1.450. This shows that there were doubts if income would affect adherence to Covid-19 preventive measures.

### Education

Education scored a mean of 2.955 and standard deviation of 1.478. This indicates that there was no assurance that the level of education affected adherence to Covid-19 preventive measures.

### Possession of Family

Possession of family scored a mean of 2.929 and standard deviation of 1.464. This means respondents were not sure if possession of family affected adherence to Covid-19 preventive measures.

### Qualitative Results

Interviews with several leaders were used to tell the importance of personal factors in explaining adoption of Covid 19 preventive measures. The views obtained are presented below.

#### a) Age

It was noted that the young people were the only group that did not take things seriously. This was caused by the fact that then only elderly people died or got critically ill. For the old people, 30 and above, adoption was very good. This was the general opinion given by all leaders interviewed.

#### b) Sex

On sex, it was revealed that both men and women respected the preventive measures. However, for male Muslims, putting on masks was a little challenging especially for those with beards which they explained as a “sunna” or a religious virtue to have long beards. The religious leaders on their part noted that,

> “*The holy books tell us to serve God but to do it wisely” (RL2, Male, 51-55, Muslim)*

#### c) Income

It was believed that income was important but even the poor could follow the preventive measures. This was the general opinion. Specifically, leaders noted that, people were fairly earing money that would serve family needs and also buy gadgets like, masks, soap and so on. One of the female leaders noted,

> *“Most of the people here are neither poor nor rich but at least can meet their livelihoods. As such, they can buy requirements for adopting SOPs in relation to Covid-19” (PL3, Female, 36-40)*

#### d) Education

The general opinion by most leaders was that education matters. However, there was no clear yardstick to tell which level of education matters. Some leaders noted that too much education is at times dangerous as it causes confusion yet little education is also a problem. One of the opinion leaders noted,

> *“in my community, it’s a mixture, you find educated people complying, but still you find others referring to so many other articles online and not complying, however, we are better off with an educated community.” (OP1, Male, 51-55)*

#### e) Possession of Family

It was generally agreed that members of society who possessed a family or who had dependents for that matter looked at preventive measures as a means to help them continue to play that bread winning role. As such, they took all measures more seriously than those who never had such responsibilities. Therefore, possession of a family encouraged adoption and adherence to the Covid-19 preventive measures.

In the correlation model above, results show that individual factors are positively and significantly correlated with adoption (r=0.412, P< 0.01). This means that a significant improvement in the individual factors will lead to significant improvements in adoption. With the exception of age (-.017), all the other factors that is sex (.160**), income (.229**), education (.285**), possessing a family (.428**) are all positively and significantly correlated with adoption. This means that improvements in income, education and possessing a family are likely to lead to better adoption of Covid-19 preventive measures.

In the regression model above, individual factors predict variance in adoption (F=80.032, Sig=.000^b^). Individual factors predict 16.7% of the variance in adoption of Covid-19 prevention measures (Adjusted R Square=.167, p<0.01). This presents individual factors as significant predictors of the variance in adoption of Covid-19 prevention measures in Entebbe. From the beta scores (Beta=.412, p<0.01) this means that a unit change in individual scores leads to a 0.412 change in adoption of Covid-19 prevention measures in Entebbe.

### 4.5 Community Factors Affecting Adoption of COVID-19 Prevention Measures among Community Members in Entebbe Municipal Council, Uganda

The results obtained are presented in table 8 below.

**Table 8:** Correlation between Community Factors and Adoption of COVID-19 Prevention Measures among Community Members in Entebbe Municipal Council, Uganda.

| **Correlations** |  |  |  |  |  |  |  |
| --- | --- | --- | --- | --- | --- | --- | --- |
|  | 1 | 2 | 3 | 4 | 5 | 6 | 7 |
| Sensitization modes (1) | 1 |  |  |  |  |  |  |
| Settlement patterns (2) | .284** | 1 |  |  |  |  |  |
| Peer influence (3) | .323** | .309** | 1 |  |  |  |  |
| Safety of face masks (4) | .227** | .240** | .310** | 1 |  |  |  |
| Risk perceptions (5) | .269** | .184** | .268** | .266** | 1 |  |  |
| Community factors (6) | .641** | .620** | .652** | .608** | .618** | 1 |  |
| Adoption (7) | .485** | .339** | .396** | .371** | .337** | .590** | 1 |
| **. Correlation is significant at the 0.01 level (2-tailed). |  |  |  |  |  |  |  |

### Community Factors Affecting Adoption of COVID-19 Prevention Measures

From table 8, overall, the results indicate that the mean score was 2.933 and standard deviation is 1.517. This means that there was no certainty on which community factors affect the level of adoption COVID-19 Prevention Measures.

### Sensitization modes

Sensitization modes scored mean of 2.864 and standard of 1.518. This shows uncertainty with high levels of divergence in opinions.

### Settlement pattern

Settlement pattern scored mean of 2.921 and standard of 1.492. This also shows that settlement patterns were doubted as to whether they affected adoption of COVID-19 prevention measures.

### Peer influence

Peer influence scored mean of 2.893 and standard of 1.504. This shows that respondents were not sure of the influence of peer influence on adoption of COVID-19 prevention Measures.

### Perceptions

Perceptions 2.892 standard of 1.505. This shows that perceptions were not clearly identified as influencing adoption of COVID-19 prevention measures.

### Safety of face masks

Safety of face masks scored mean of 2.902 standard of 1.505. This shows that there was no certainty on the use of face masks in COVID-19 prevention.

### Risk perceptions

Risk perceptions scored mean of 2.902 and standard of 1.501. Risk perception results show that respondents were not sure on how risk perceptions affected adoption of COVID-19 Prevention Measures.

### Qualitative Results

The study also obtained results on community factors that could be affecting adoption of Covid-19 preventive measures. The responses obtained are presented below.

#### a) Sensitization modes

Leaders agreed that sensitization matters. It as reasoned that Covid-19 was new to everyone including those who thought were experts. It was thus acknowledged that sensitization was important. One of the leaders noted that, he had his community get sensitized by health teams and he was seeing results already,

> “*Previously, I was very green and so were my people about Covid. However, when I linked with health workers and they accepted to sensitize people, I inclusive, things changed for the better. People now know why and how when it comes to Covid-19 prevention. It is only those who stubbornly dodged that are still disturbing us” (PL2, Male, 51-55).*

Another leader however was not happy that the community is not sensitized enough, and this explained low adoption of SOPs,

> *“My area, apart from TV adverts, people have just to follow what we tell them, otherwise, they know little. You can imagine, some up to now do not know how to properly wear a mask. Government or the ministry of health needs to do more here”PL3, Female, 36-40)*.

From the above, there were gaps in sensitization much as it was noted to be a key enabler of adoption.

#### b) Settlement pattern

Leaders acknowledged that Entebbe on the face is good looking but inside, there are slums like Lugonjo where people leave in congestion. This defeats adoption of social distancing and even hygiene in such areas is compromised. Still, being in a slum prevents health workers from easily reaching out to such areas. Leaders largely noted that the settlement patterns were a hindrance to some citizens to adopt the measures. Actually, even the vaccination teams were reported to distance themselves from slums citing perceived insecurity.

#### c) Peer influence

Apart from the youths, peer influence was not so pronounced in Entebbe according to almost all the interviewees. Positive peer influence was also reported among people sharing the same trade as boda-boda cyclists, market vendors, saloon, and hairdressers among others.

#### d) Perceptions

The general perceptions were mixed according to leaders. The biggest challenge was that Covid-19 coincided with the political season. This raised mixed feelings. However, after elections and following death of people known to the locals, the perceptions quickly changed, and adoption was faster and deeper.

#### e) Safety of face masks

According to the leaders interviewed, face masks were largely locally made, and some were from tailors known to the intended users. This improved the perception that the masks were safe to use. With only exceptions being people with breathing difficulties due to lung related infections like TB and Asthma.

#### f) Risk perceptions

Covid-19 by the time the study was done had killed several people known to the residents. This was enough to make them appreciate that it was real and risky to get into contact with Covid-19. This made adoption of preventive measures the only choice left.

From table 11, results indicate that there is a positive, significant, and moderate effect of community factors on adoption of COVID-19 Prevention Measures (r=.590^**, P^< 0.01). This means that positive changes in community factors are likely to lead to positive changes in adoption of COVID-19 Prevention measures in Entebbe since the two variables exhibit positive correlation. All the community factors show positive and significant associations with adoption of COVID-19 prevention measures, that is, Sensitization modes (.485**); Settlement patterns (.339**); Peer influence (.396**); Safety of face masks (.371**); and Risk perceptions (.337**). This means that improvements in all these factors is likely to lead to better adoption of Covid-19 preventions measures.

From the regression model in table 10 above, result confirm that community factors predict adoption of Covid-19 preventive measures (F=209.202, Sig.= .000^b^). Community factors predict 34.6% of the variance in adoption of Covid-19 preventive measures (Adjusted R Square=.346, P< 0.01). This means that community factors once improved, there is likely to be better adoption of Covid-9 preventive measures. From the beta scores (Beta= .590, p< 0.01), results show that a unit change in community factors leads to a 0.590 change in adoption of Covid-19 preventive measures. This confirms that enhanced community factors are likely to be followed by improved adoption of Covid-19 preventive measures in in Entebbe Municipal Council, Uganda.

**Table 9:**
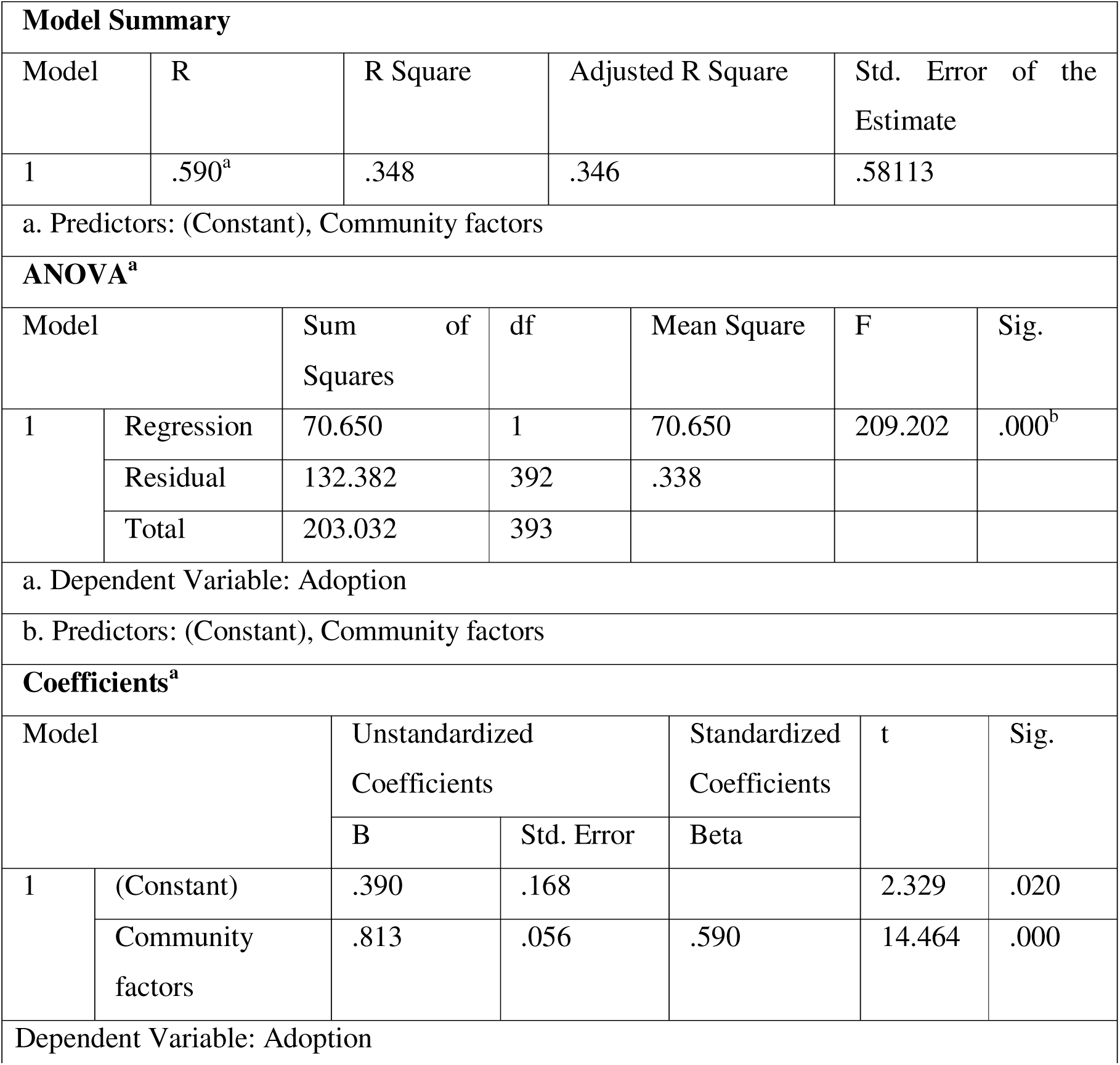
Regression between Community Factors and Adoption of COVID-19 Prevention Measures among Community Members in Entebbe Municipal Council, Uganda.

**Table 10:**
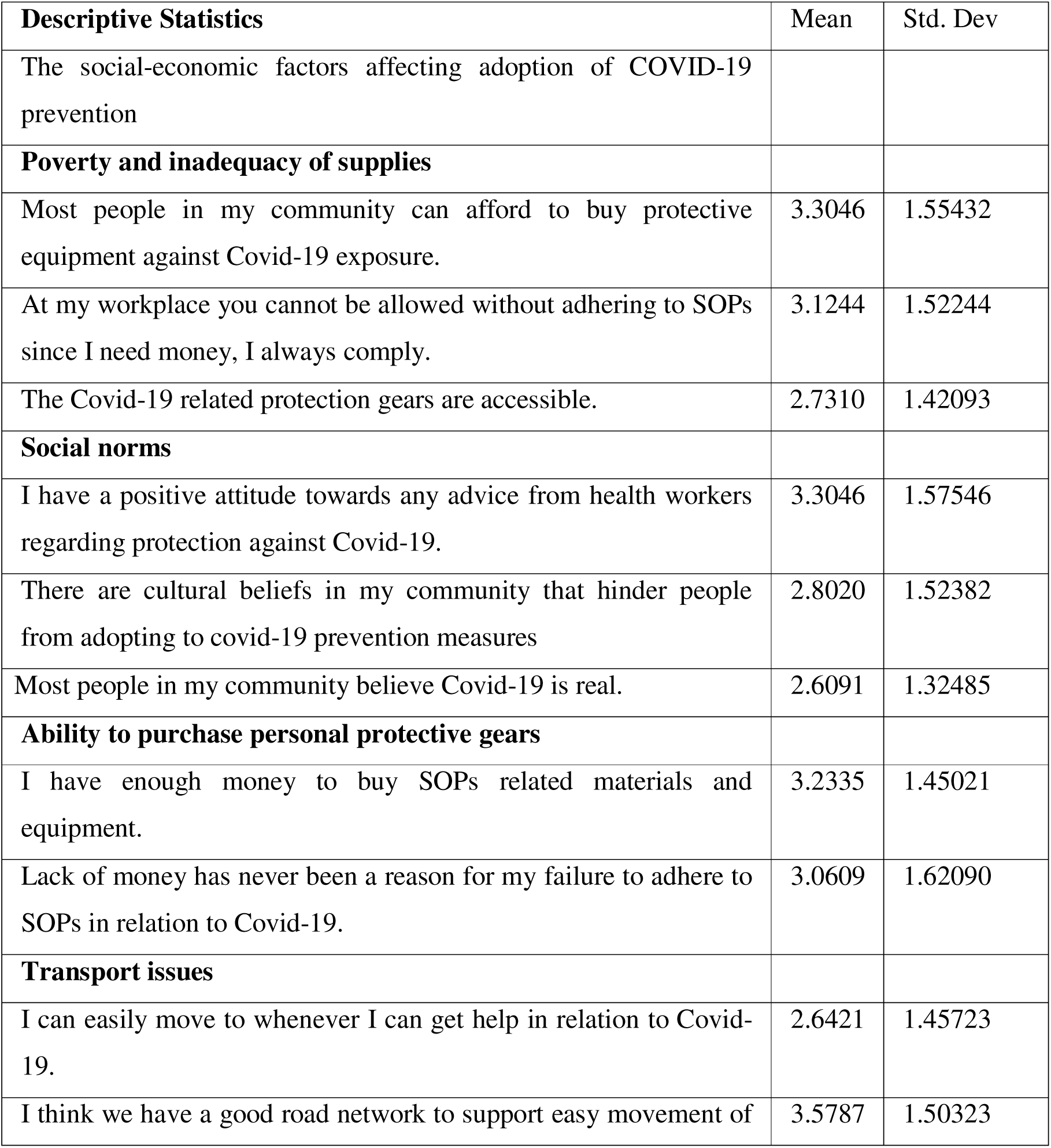
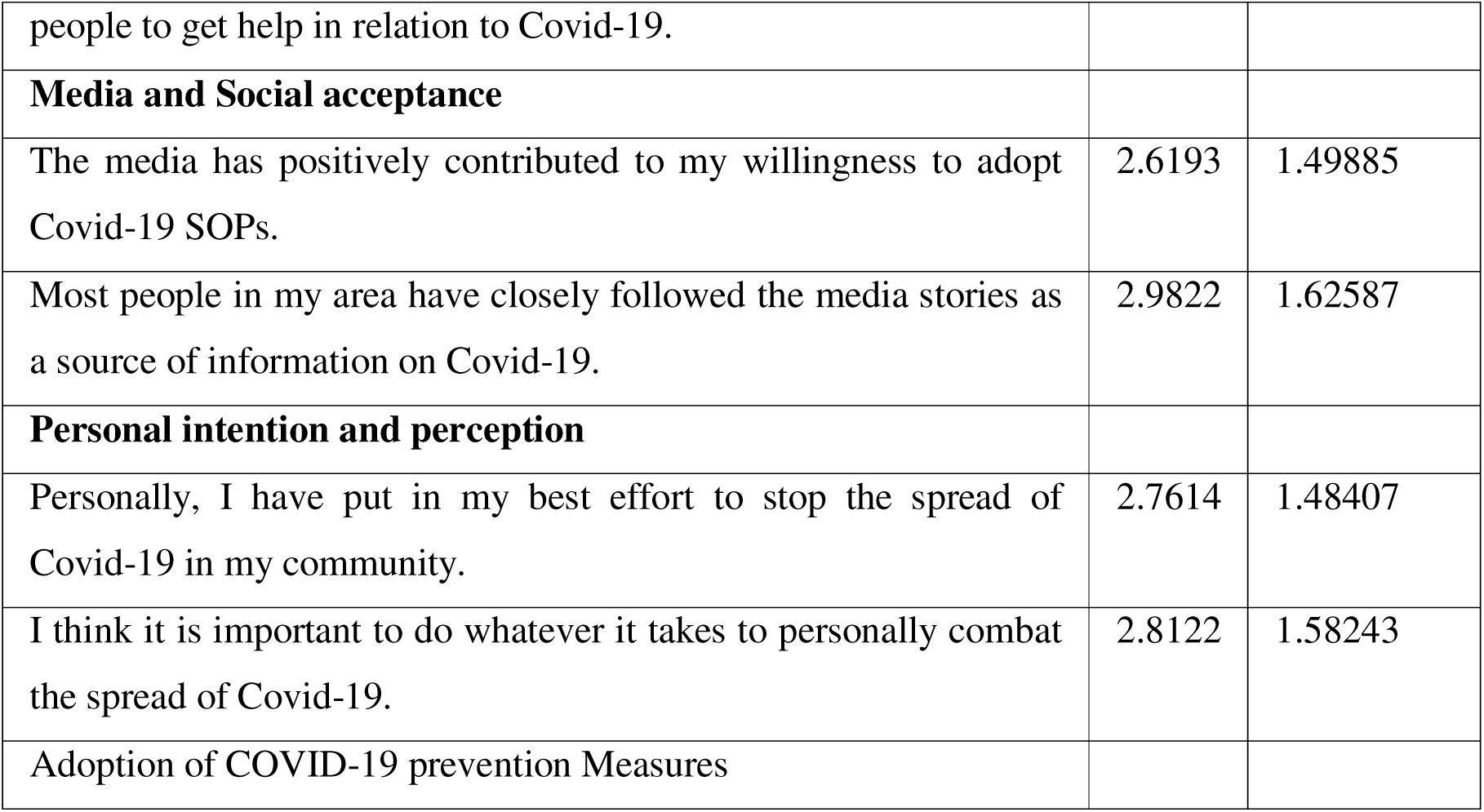
Social-economic factors affecting adoption of COVID-19 prevention measures.

### Social-economic factors affecting adoption of COVID-19 prevention measures among community members in Entebbe Municipal Council, Uganda

The results obtained are presented in table 11 below.

**Table 11:** Correlation between Social Economic Factors and Adoption of COVID-19 Prevention Measures among Community Members in Entebbe Municipal Council, Uganda.

| **Correlations** |  |  |  |  |  |  |  |  |
| --- | --- | --- | --- | --- | --- | --- | --- | --- |
|  | 1 | 2 | 3 | 4 | 5 | 6 | 7 | 8 |
| Poverty and inadequacy of supplies (1) | 1 |  |  |  |  |  |  |  |
| Social norms (2) | .250** | 1 |  |  |  |  |  |  |
| Ability to purchase personal protective gears (3) | .227** | .289** | 1 |  |  |  |  |  |
| Transport issues (4) | .213** | .326** | .144** | 1 |  |  |  |  |
| Media and Social acceptance (5) | .186** | .366** | .344** | .174** | 1 |  |  |  |
| Personal intention and perception (6) | .305** | .386** | .254** | .291** | .257** | 1 |  |  |
| Social economic factors (7) | .615** | .704** | .594** | .553** | .617** | .658** | 1 |  |
| Adoption (8) | .256** | .446** | .275** | .302** | .502** | .489** | .602** | 1 |
| **. Correlation is significant at the 0.01 level (2-tailed). |  |  |  |  |  |  |  |  |

### Overall score on Social-economic factors

Results show an overall mean of 2.957 and standard deviation of 1.512. This means that there was no certainty as to how the community factors influence adoption of COVID-19 prevention measures in Entebbe.

### Poverty and inadequacy of supplies

Poverty and inadequacy of supplies exhibited a mean 3.053 and standard deviation of 1.499. This means respondents were not sure if supplies and poverty affected adoption of Covid-19 preventive measures. This was obtained however with a high standard deviation thus sharp differences in opinions on the same.

### Social norms

Social norms indicate a mean of 2.905 and standard deviation of 1.475. This result casts doubts on the effect of social norms on adoption of Covid-19 preventive measures. The high standard deviation means sharp differences in opinions on the same.

### Ability to purchase personal protective gears

Ability to purchase personal protective gears revealed and average score of 2.979 and standard deviation of 1.487. This means not sure and thus respondents were not certain if ability to purchase protective gears explained adoption levels.

### Transport issues

Transport issues shows a mean of 2.942 and standard deviation of 1.481. This shows uncertain but with a high standard deviation meaning that transport issues were accepted as affecting adoption of the Covid-19 preventive measures but with hesitation, hence the large standard deviation.

### Media and Social acceptance

Media and Social acceptance indicated a mean of 2.961 and standard deviation of 1.484. This shows that partly social media and social acceptance affects adoption. The high standard deviation however casts doubt for many hence uncertainty being the overall finding here.

### Personal intention and perception

Personal intention and perception scored a mean of 2.952 and standard deviation of 1.482. Results also confirm that there was uncertainty on whether personal intention and perception affected adoption of Covid-19 preventive measures.

### Qualitative Results

The study in addition to the quantitative results above also gathered qualitative information on the social economic factors and how they influenced adopt of covid-19 preventive measures as presented below in the different sections which also double as the themes identified.

#### a) Poverty and ability to purchase personal protective gears

Leaders noted that what matters is not the ability, instead, it is the willingness. Whereas SoPs called for some small spending, there are people who never cared to buy a mask of 1000 shillings and could afford a meal of 7,000 in a restaurant. As indicated by one of the opinion leaders,

> *“I see relatively poor pole carrying even a spare mask. At the same time, I see people driving but with no mask. Yes, money is important, but the will supersedes.” (OP1Male, 51-55)*

This was supported by a religious leader who noted,

> *“my brother, let me tell you, even here in Mosques, it’s about the heart to give not the wealth one has. This too applies to Covid, there are those who can even afford to donate covid-19 preventive equipment’s like masks, sanitizers, soap etc. to the whole zone but you find them not using them. Can we say they can’t afford, No. it’s a perception issue.” RL2Male, 51-55, Muslim*

#### b) Transport issues

In Entebbe there was good transport day and night and members of parliament had given out many ambulances in addition to private and government ambulances. So transport for example of suspects to facilities to prevent spread of the diseases was never a problem. However, in case of late-night emergencies, some challenges related to transport were reported but these were quickly solved but linking with local leadership. Thus, transport is a key enabler but in Entebbe, it was reported to be largely good enough.

#### c) Media and Social acceptance

The media was seen as both positive and negative. All leaders agreed that we cannot go back to government monopoly of the media but there was need to improve on the regulation of what was being broadcast and printed in the media papers. Leaders agreed that media is a very strong influencer, and it ought to be used cautiously not for Covid-19 only but for any public health issues.

#### d) Personal intention and perception

Personal intention as it was not commented on much by leaders. However, majority noted that it was important to ensure that people make decisions not out of excitement or frustration but out of information and right perception. Some of the negative perceptions related to safety of the vaccines, whether Covid-19 was manufactured and the intention and if we need to trust those who allegedly manufactured it to give use free donations of masks and vaccines and so on. Such ideas were noted to be baseless but an indicator that the wrong perceptions were yet to be driven out through mass sensitization.

Results from table 12 confirm that there is a positive, significant, and high correlation between social-economic factors and adoption of Covid-19 preventive measures (r=.602^**,^ p<0.01). This means that improvements in the economic factors are likely to lead to better adoption of Covid-19 preventive measures. Also, the elements of social-economic factors exhibited positive and significant associations with adoption of Covid-19 measures; poverty and inadequacy of supplies (.256**); social norms (.446**); ability to purchase personal protective gears (.275**); transport issues (.302**); media and social acceptance (.502**) and personal intention and perception (.489**).

**Table 12:** Regression between Social Economic Factors and Adoption of COVID-19 Prevention Measures among Community Members in Entebbe Municipal Council, Uganda.

| Model Summary |  |  |  |  |  |  |
| --- | --- | --- | --- | --- | --- | --- |
| Model | R | R Square | Adjusted R Square | Std. Error of the Estimate |  |  |
| 1 | .602<sup>a</sup> | .363 | .361 | .57441 |  |  |
| a. Predictors: (Constant), social economic factors |  |  |  |  |  |  |
| ANOVA<sup>a</sup> |  |  |  |  |  |  |
| Model |  | Sum of Squares | df | Mean Square | F | Sig. |
| 1 | Regression | 73.691 | 1 | 73.691 | 223.337 | .000<sup>b</sup> |
|  | Residual | 129.341 | 392 | .330 |  |  |
|  | Total | 203.032 | 393 |  |  |  |
| a. Dependent Variable: adoption |  |  |  |  |  |  |
| b. Predictors: (Constant), social economic factors |  |  |  |  |  |  |
| Coefficients<sup>a</sup> |  |  |  |  |  |  |
| Model |  | Unstandardized Coefficients |  | Standardized Coefficients | t | Sig. |
|  |  | B | Std. Error | Beta |  |  |
| 1 | (Constant) | .725 | .140 |  | 5.166 | .000 |
|  | Social economic factors | .691 | .046 | .602 | 14.944 | .000 |
| a. Dependent Variable: adoption |  |  |  |  |  |  |

From the above regression model, results show that social-economic factors predict adoption of Covid-19 preventive measures (F=223.337, Sig.= .000^b^). Also, the social economic factors together predict 36.1% of the variance in adoption of Covid-19 measures in Entebbe (Adjusted R Square=.361, P<0.01). This means that changes in the social =economic factors are likely to lead to significant changes in adoption of Covid-19 preventive measures. Looking at the beta (Beta=.602, P<0.01) reveals that a unit change in social –economic factors will lead to a 0.602 change in adoption of Covid-19 preventive measures.

## Discussion

### Individual factors affecting the adoption of COVID-19 prevention measures

Overall, individual factors predict 16.7% of the variance in adoption of Covid-19 prevention measures. The study explored individual factors affecting adoption of Covid-19 Prevention Measures among Community Members in Entebbe Municipal Council, Uganda. Overall, there was no certainty on how individual factors influence adoption of Covid-19 prevention measures. Results show that individual factors are positively and significantly correlated with adoption. A significant improvement in the individual factors will lead to significant improvements in adoption. With the exception of age, all the other factors that is sex, income, education, possessing a family all positively contribute to adoption. It is also confirmed that individual factors predict variance in adoption and are significant predictors of the variance in adoption of Covid-19 prevention measures in Entebbe.

### Discussion

Several individual factors have been reported to influence adoption of COVID-19 preventive measures as discussed below.

### Age

The study revealed a negative correlation between age and adherence (-.017). This means that increasing age led to decrease in adherence. In the current study age seems not to be an important determinant of adoption of preventive measures. This agrees with (Jordan, Adab and Cheng, 2020; Zhou *et al*., 2020) however, the results contrast with (Felsenstein and Hedrich, 2020; Plaza-Zamora *et al*., 2020; Feyisa, 2021). The study has revealed that old people were given scaring information that Covid-19 targets the old people and indeed most of the deaths occurred among those aged 55 and above according to most respondents in the interviews. The general perception thus is that old people at a bigger risk compared to younger ones hence creating a scenario of forced adherence to preventive measures. To policy makers, this study suggest that those tasked with ensuring compliance ought to look out for what fits every age category to enhance compliance.

### Sex

The study revealed a positive and significant relationship between sex and adherence (.160**). This ideally means that there are differences in adherence with females being more compliant. Sex has been revealed to have positive associations with adoption of Covid-19 preventive measures. It was however revealed in interviews that women tend to be more careful, critical and honesty in adherence to measures including public health. As such, they were revealed to be more willing to adopt the Covid-19 preventive measures. Women were mostly at home or nearby thus helping them to be strict and besides were shouldering most of the burden of caring for the sick and looked ad adopting preventive measures as directly relieving them of the duties to look after the sick which would follow if they never adhered to the measures in place. These results seem to agree with (Galasso *et al*., 2020; Garikipati, 2020; Feyisa, 2021; Padidar *et al*., 2021). All these studies have reported comparatively better adherence of Women compared to men when it comes to public health and other health directives and measures. The results however contrast with (Chen *et al*., 2014) where the findings revealed that male patients adhere more effectively to medications than female patients do. Overall, women have proved to be more reliable when it comes to adoption of preventive health measures. This challenges the male counterparts to improve on this for better general health thus future interventions need to look into this closely. As such, the fact that men are less inclined to adoption of public health measures calls for special efforts to attract men to be part of public health interventions.

### Income

The study revealed that income (.229**) was positively and significantly related to adherence. In the current exploration, results have revealed that income is important in improving adoption. This is supported by the fact that all gadgets including face masks, washing soap and water, even maintaining a social distance call for parting with some money. It is not however clear; how much income would be a good motivator of adoption. Still, in the interviews, there were cases of the somehow well off people who had all the abilities to buy all that is needed for adoption of preventive measures for Covid-19, but they stubbornly never cared. Overall, income is important as per the current study and this has been supported partly by (Padidar *et al*., 2021) and partly by (Odoi, 2020) who report that livelihoods and the need to make a living were the most common explanatory factors. The final position of the study is that income is an important enabler but does not guarantee adoption of preventive measures. It is thus asserted that higher income levels are desirable but not conditional when it comes to adoption of public health and preventive measures. It is thus imperative that poverty is fought using multi-dimensional approach for better results.

### Education

The study revealed that education (.285**) was positively and significantly related to adherence, a finding which agrees with (Shi *et al*., 2020). It has been confirmed that people need to be given information, but in the right form, level of depth and with a good packaging to stimulate adoption. The study has revealed information under load and overload as being undesirable extremes that prevent adoption. People with too much information got confused and those with little information were not sure why and whether. It is thus important to have people well informed in order to foster adoption of health directives and preventive measures. These findings relate with literature, for instance (Plaza-Zamora *et al*., 2020) and (Shi *et al*., 2020) note that education level and professional training are very instrumental in shaping people’s knowledge, attitudes, and practices regarding COVID-19. Similarly, (Pinchoff and Tidwell, 2020) and (Abdulkareem *et al*., 2020) note that knowledge is crucial in shaping people’s behavior and practices especially during any disease outbreak. In the same way, knowledge level has been linked with panic emotion thus influencing attitude and practices toward COVID-19. In the same way (Zhong *et al*., 2020), highlights the threat of misinformation and fake news (Zhong *et al*., 2020) while higher education level and higher income have been linked to better adoption of preventive measures (Feyisa, 2021). Generally, education is important but more importantly, the content and how it is delivered is key.

### Possession of Family

The study revealed that possessing a family (.428**) was strongly and positively correlated with adherence. Having a family in Uganda is generally linked to being responsible, much not all those who have families are responsible. Women and men with families especially being the bread winners saw themselves as being indispensable to their families thus forcing them to adopt whatever measure that was presented to prevent Covid-19. The risk would be that if one has a family and gets infected, then the entire family was likely to get infected and this was a great motivation for adoption. Hager et al. (2020) has highlighted possession of family as being an important motivator for adoption of preventive measures and this too applies to Covid-19 preventive measures adoption.

### Community factors affecting adoption of COVID-19 prevention measures

The study revealed that there is a positive, significant, and moderate effect of community factors on adoption of COVID-19 Prevention Measures (r=.590). The study also explored the community factors affecting adoption of COVID-19 prevention measures. The study has confirmed a positive, significant, and moderate effect of community factors on adoption of COVID-19 Prevention Measures. The study confirms that positive changes in community factors are likely to lead to positive changes in adoption of COVID-19 Prevention measures in Entebbe since the two variables exhibit positive correlation. All the community factors show positive and significant associations with adoption of COVID-19 prevention measures, that is, Sensitization modes, Settlement patterns, Peer influence, Safety of face masks and Risk perceptions. This means that improvements in all these factors is likely to lead to better adoption of Covid-19 preventions measures. The study also confirms that community factors predict adoption of Covid-19 preventive measures and once these factors once improved, there is likely to be better adoption of Covid-9 preventive measures.

### Discussion

Several community factors have been reported to influence adoption of COVID-19 preventive measures as discussed below.

### Sensitization modes

The study revealed that sensitization modes (.485**) were positively and significantly correlated with adherence. The study has confirmed that sensitization modes are important in influencing adoption of preventive measures. It was highlighted that the use of electronic media like social media, TV, non-electronic media like radios, newspapers, community public address and having a sensitization strategy were important factors. These findings are supported by literature where it is noted that the way the public gets the information about covid-19 SOPs could be determining their response by complying or not (Ikoja-Odongo, 2002). This position is however nullified by the feeling that Covid-19 has been recently used for political gains (Odoi, 2020). Nevertheless, it is important to use believable, accessible, and sustainable means of delivering information. The intentions of the deliverer of the information should also be clear and honest to improve acceptance and adoption of the preventive measures as seen in Uganda’s case based on evidence from Entebbe.

### Settlement pattern

In addition, the study revealed that the settlement patterns (.339**) are positively and significantly correlated. The study also confirms that settlement pattern is an important issue in influencing adoption of preventive measures. Being in a crowded place, having adequate rooms interfered with adherence to SOPs like social distancing. The study confirms that the settlement patterns are vital in determining adoption of preventive as they influence what is possible, accessible, and affordable. These findings have been supported by literature where it is noted that perceived susceptibility to COVID-19 was higher among people who are living in crowded places compared to those who stayed in less crowded places (Mya *et al*., 2020). It was further noted that living arrangements in the informal settlements affect people’s adherence to SOPs like social (Sim, Moey and Tan, 2014; Kasozi *et al*., 2020; Odoi, 2020; unicef, 2020) distancing reveal that living in Kampala City Centre was associated with high adherence to preventive measures. Furthermore, living in a household with other siblings is less likely to cause adherence to the preventive measures (Kasozi *et al*., 2020). It is thus noted that even when someone is willing to adopt preventive measures, the place of residence, the size of the rooms, the distance from home to home, the number of occupants all can support or interfere with level of adoption of the Covid-19 preventive measures.

### Peer influence

The study further revealed that peer influence (.396**) is significantly and positively correlated with adherence. Peer influence is common regardless of age, sex, income, occupation and so on. However, in the current study, it has been common in the youth where the community tends to follow their peers who do not adhere to the good practices. It has also been singled out that members who share common trade for example market workers, taxi drivers tend to be influenced by each other. It is also noted that people who abuse drugs tend to influence people near them not to adhere to SOPs. This actually explains the fact that bars were among the last places to have the operation ban lifted. These findings are in line with literature where it is reported that peer influence if negative is worsened by the limited sensitization to the community (Feyisa, 2021). It is also noted that youths are 93% times less likely to adhere to preventive measures, influenced by peer influence (Feyisa, 2021). Also, people tend to believe more in their peers than any other influencer as revealed by Ahmad et al. (2021). From this, it is clear that peer influence has significant impact and if positive, can be a good supportive tool for adoption.

### Perceptions

Risk perceptions is positively and significantly correlated with adherence (.337**). People have always been with different perceptions, and these significantly influence adoption of the preventive measures. Such perceptions, negative, positive, true, and false always influence acceptance or rejection of whatever measures or directive including Covid-19 preventive measures. It was specifically noted that people who have not lost close friends or relatives took SOPs more seriously. This resonates properly with literature where it is said that people protect themselves based on four factors to the current threat: the perceived severity of a threatening event, the perceived probability of the occurrence, or vulnerability, the efficacy of the recommended preventive behavior, and the perceived self-efficacy. In addition, the level of risk perception is affected by an individual’s exposure to different sources of information (Kwok, K. O., Li, K. K., Chan, H. H., Yi, Y. Y., Tang, A., Wei, W. I., & Wong, 2020). (Honarvar *et al*., 2020) in relation to the current study submits that community responses are important for outbreak management during the early phase of the pandemic. It is also reported that people with poor knowledge; attitude and practice towards COVID-19 prevention were more affected by COVID-19 (Cucinotta and Vanelli, 2020). All these illustrate how important perceptions are in determining adoption of Covid-19 preventive measures.

### Safety of face masks

Regarding safety of face masks, a positive and significant relationship (.371**) was noted with adherence to preventive measures. Safety of face masks has been proved to have significant links with adoption of preventive measures. It has been revealed that the general feeling is that wearing face masks was important and that most of the face masks in place were effective. This kind of reasoning was important and improved adoption. However, there were a section of people who opposed safety of face masks. First, that the masks were substandard, and that Covid-19 makes breathing difficult and that putting on masks would weaken the lungs before the Covid-19 attack and that if the attack happens, then the lungs would be too weak. This misconception was actually common but based on rumors with no scientific proof. This was thus a knowledge gap that calls for attention. The popularity of face masks however is questioned due to some features as noted by (Dohrn *et al*., 2022) that discomfort resulted from poor fit of the facemasks, which was due to certain facial structures of the wearer. In addition, facemasks tend to become damp after a period of time in warm environments, further contributing to the discomfort of mask-wearing (Hallock, 2020). However, the use of face masks has been found to be popular as noted by (Purens, 2020) and (Abdullahi *et al*., 2020).

### Social-economic factors affecting adoption of COVID-19 prevention measures

Overall, the study revealed significant and high correlation between social-economic factors and adoption of Covid-19 preventive measures (602**). The study also explored the social-economic factors affecting adoption of COVID-19 prevention measures. The study revealed is a positive, significant, and high correlation between social –economic factors and adoption of Covid-19 preventive measures. As such, improvements in the social –economic factors are likely to lead to better adoption of Covid-19 preventive measures. The elements of social-economic factors exhibited positive and significant associations with adoption of Covid-19 measures; poverty and inadequacy of supplies, social norms, ability to purchase personal protective gears, transport issues, media and social acceptance and personal intention and perception. Social-economic factors predict adoption of Covid-19 preventive measures. Changes in the social economic factors are likely to lead to significant changes in adoption of Covid-19 preventive measures.

### Discussion

Several social-economic factors have been reported to influence adoption of Covid-19 preventive measures as discussed below.

### Poverty and inadequacy of supplies

The revealed that poverty and inadequacy of supplies is positively and significantly correlated with adherence (.256**). The current study shows that poverty is not a big hindrance as supplies are noted to be available at different prices with masks costing as low as Uganda shillings 500 to Uganda shillings 15000, sanitizers costing Uganda shillings 3000 up to Uganda shillings 50,000 and so on. It was also noted that at most workplaces, employers were not allowing in workers without masks and so on. Since money was needed, the workers had no choice but to comply. Also, there was a good level of accessibility to the Covid-19 related protection gears. These results are in line with literature where it is stated that resistance and poor socioeconomic status encourage transmission of COVID-19 in its infancy stage (Abdelhafiz *et al*., 2020). In addition, higher socio-economic status and higher educational attainment are often related to engagement in better behaviors (Wirz et al., 2020). In the same way, the risk of COVID-19 is more significant for those that live in Low-and Middle-Income Countries, where Uganda is one of them, as well as rural, and poverty-stricken areas (Waterfield *et al*., 2021). However, this assertion cannot be justified today as Covid-19 has hit the more developed economic harder. Some only argue that the victims in Africa are only low because of poor reporting systems. Thus, the question of social economic status affecting adoption, statistically is significant but still their issues that have be dug deeper to get better insights.

### Social norms

In addition, the study revealed that social norms (.446**) were positively and significantly correlated with adherence. Culture influences general behavior and is such a significant influence. However, since the study had many tribes with no particular dominant culture since tribe and culture were diffused. For instance, many people from across all the tribes spoke Luganda and behaved and believed almost like Baganda and Banyankore since they grew up in similar environments. What existed was thus a myth connected to particular sections of society. This has also been confirmed by the fact that some cultural beliefs in the community hindered people from adopting to covid-19 prevention measures for instance, Muslims with beards found it hard to wear masks but it was so easy to do hand washing since it was already part of their norms. Most people in the community believed Covid-19 was real but some trusted that the ancestral spirits would take control. Some sections were opposed to alcohol and thus saw the use of sanitizers as leaning towards alcohol. This makes it important to consider the cultural implications of health interventions. These findings are in line with literature where it is indicated that social norms are one of the factors that affect utilization of preventive measures (Bronfman *et al*., 2021). It is further asserted that favorable attitude, combined with the approval of the behavior by others close to them and a positive perception of control strengthens the intention to perform that particular behavior (Ajzen, 2020).

### Ability to purchase personal protective gears

The study further noted that ability to purchase personal protective gears (.275**) was positively and significantly correlated with adherence to the protective measures against Covid-19. The study confirms a strong association between ability to purchase personal protective gears and adoption of the preventive measures. It was noted that availability of money to buy SOPs related materials and equipment was an important issue. However, lack of money was rarely a reason for failure to adhere to SOPs in relation to Covid-19. This is supported by the fact that not all SOPS demanded parting with money, for instance, social-distancing, vaccination, washing hands all involved negligible spending if any. This issues of affordability have been reflected earlier that facemasks affordability has been reported to affect compliance with these health behaviors and that the cost of preventive measures tends to shoot to the highest once the pandemic is in initial stages. This reasoning explains why today everything related to Covid-19 is becoming more affordable since the initial stagers have been crossed.

### Transport issues

Regarding transport issues, a positive and significant correlation was found with adherence (.302**). It was further noted that transport was an important influencer but was in a good state as it was available and cheap. It was indicated that people could easily move to whenever they could get help in relation to Covid-19. Ambulances were in place and the political leaders offered transport at no or relatively very little charges (fuel only). However, without transport, then congestion in the limited number of vehicles easily undermined social distancing. Still, getting medical attention for emergencies late night was problematic for those who never had access to vehicles or available means of transport. It has been pointed out that people who often mix-up with the community tend to be better compliant (Ear, 2012).

### Media and Social acceptance

It was revealed that media and social acceptance (.502**) was positively and significantly correlated with adherence. Uganda is well enriched in terms of variety of media, thus making it a positive and significant influencer of adoption of Covid-19 preventive measures. The media is seen to positively contribute to willingness to adopt Covid-19 SOPs. Most people closely followed the media stories as a source of information on Covid-19. However, given the liberty to publish, some medical content was exaggerated and misleading. This created negativities which required immediate correction before creating image damage of the efforts to combat Covid-19. Reports such as Covid-19 kills on the elderly, Covid-19 affects more males than females, vaccination of Covid-19 is a slow but fatal poison, causing even infertility and low sexual libido were common. With the good efforts of the ministry of health and responsible media, there were finally counteracted with positive messages. It was earlier noted that pressure from the media, in the form of impactful advertising campaigns has been shown to contribute to the high levels of adherence to mask-wearing in Japan (Charmes, 2020). The role of social media and its acceptance features in (Chew *et al*., 2020).

### Personal intention and perception

The study revealed that personal intention and perception (.489**) was significantly and correlated with adherence. The study presents personal intention and perceptions as significant contributors to adoption of Covid-19 preventive measures. The study indicates people putting in personal efforts to campaign for adoption of preventive measures. There was also a strong commitment by some sections of society to take personal initiatives, without pay or position as leaders but being health nationalists to do whatever it takes to personally combat the spread of Covid-19. Such people donated face masks, encouraged vaccination, donated hand washing facilities for communal use and sharing any correct information they knew about preventing the spread of Covid-19. It is thus revealed that sensitization is one of key use of social media and public figures who can attract good voluntary following (Charmes, 2020; Hills and Eraso, 2021). However, forceful implementation is seen to cause positive impact on adoption but which is short lived as it is not based on personal conviction and thus not sustainable.

### Conclusion

Personal factors significantly influence adoption of preventive measures. Improvements in such factors as income and education is vital in the efforts to improve Covid-19 preventive measures adoption. However, the difference between women and men’s adoption, with women being better adopters call for attention just like the young people’s negligence to adoption of preventive measures.

Pandemics affect communities collectively and the role of community factors is worth attention. Collectively, important factors have been identified to comprise in order of rank, sensitization modes, peer influence, perceived safety of face masks, settlement patterns and risk perceptions. Once these factors are in a good state, the adoption probabilities are likely to improve.

Collectively and in comparison, with individual and community factors, social-economic factors are presented as the greatest influencers of adoption of Covid-19 preventive measures. Looking the individual elements reveals in the order of rank the most outstanding factors to constitute; media and social acceptance, personal intention and perception, social norms, transport issues, ability to purchase personal protective gears and poverty and inadequacy of supplies.

## Data Availability

All data produced in the present work are contained in the manuscript

## Acknowledgements

We take this opportunity to express a deep sense of gratitude to lecturers at Uganda Martyrs University School of public health, parents and to all classmates and employers for their invaluable information and grateful for their cooperation during my study period.

We would also like to acknowledge the participation of the residents of Entebbe Municipality for providing the primary data.

## Notes

### Competing Interest Statement

The authors have declared no competing interest.

### Funding Statement

This study did not receive any funding

### Author Declarations

The Research Ethics Committee of The Aids Support Organisation gave ethical approval for this work

